# Dissecting the relationship between haplotypes around ATXN2 CAG repeats and the number of CAA interruptions by long-read sequencing

**DOI:** 10.64898/2026.03.11.26348169

**Authors:** Beoung Hun Lee, Joe Chan, Yuk Yee Leung, Corey T McMillan, NYGC ALS Consortium, Yuanquan Song, Defne A Amado, Kai Wang

**Author notes:** Correspondence should be addressed to KW. See Table S4 for the list of consortium members (NYGC ALS Consortium).

## Abstract

**Background:** CAG repeat expansions in ATXN2 are implicated as risk factors for several neurological diseases, including spinocerebellar ataxia type 2 (SCA2) when >=33 CAG repeats are present, and amyotrophic lateral sclerosis (ALS) when 27-33 CAG repeats are present. However, how haplotypes around the repeats and CAA interruptions within the repeats are associated with disease phenotypes remains poorly understood. Previous studies on haplotypes around ATXN2 were limited to SNPs very close to the repeats (<5kb) or were based on statistical inference only.

**Methods:** Here, we used long-read sequencing on the Oxford Nanopore Technologies (ONT) platform to simultaneously infer haplotypes around *ATXN2*, the number of CAG repeats, and the number of CAA interruptions, along with NYGC ALS Consortium NGS dataset. We further sequenced 41 individuals (EUR = 39) with neurological diseases with intermediate repeats by ONT.

**Results:** We found that haplotypes around *ATXN2* and the number of interruptions show ethnicity-specific and ALS-specific distribution. Three CAA interruptions are present at low prevalence (∼1%) in control populations in multiple ancestry groups, but high prevalence (∼55%) in ALS individuals with intermediate repeats. Furthermore, we examined 159 individuals with ALS (∼90% European ancestry) with intermediate *ATXN2* repeats and found a unique haplotype in ALS individuals with three CAA interruptions, which can be tagged by an SNV, rs148019457. We also validated that the rs148019457-G allele is only present in haplotypes with three CAA interruptions.

**Conclusions:** In summary, our study shows that 3 CAA interruptions are rarely seen in healthy controls but are common in those with expanded ATXN2 CAG repeats who have neurological disorders, and that rs148019457 tags a specific haplotype with 3 CAA interruptions within expanded ATXN2 CAG repeats in individuals of European ancestry. These results have implications for the development of precision genomic medicine for neurological disorders, and the tag SNP may help identify those with interruptions from existing population genotyping data.

## Background

Several neurological diseases such as spinocerebellar ataxia 2 (SCA2), amyotrophic lateral sclerosis (ALS), and Huntington’s disease (HD) are caused by short tandem repeat (STR) expansions. While rare themselves, these diseases in total affect millions of people worldwide and some studies suggest that many of the diseases affected by STR may be underdiagnosed[1–4]. These repeat expansions are traditionally diagnosed using targeted approaches including PCR-based methods, fragment length analysis and gel electrophoresis analysis, as the repeat sequences make it difficult to identify the number of repeats in the regions using next-generation sequencing (NGS), which produces reads that are 150-300 bps in size. Furthermore, while PCR or Southern blot can determine the number of simple repeats, these methods cannot distinguish whether the sequence that they have found contains repeat interruptions (for example, CAA sequences within CAG repeats)[3]. NGS can determine the sequence or identify interruptions if the regions are smaller than its read size, but has limitations both with longer regions and with identifying genotypes of the SNVs surrounding the repeats and the haplotypes that are associated with expanded repeats.

The more recent development and implementation of long-read sequencing like Pac-Bio and Oxford Nanopore Technology (ONT) has enabled superior diagnosis and analysis of repeat expansion diseases. Pac-Bio Sequencing utilizes SMRTbell, in which double-stranded DNA molecules with hairpin structures enter a flow cell and have fluorescent-labeled nucleotides added to identify DNA sequences, similar to Sanger sequencing[5]. ONT sequencing takes advantage of a motor protein that can measure the change in an electric current when a DNA sequence passes through its pore, and identifies DNA sequences based on the change in the electric current. Pac-Bio produces reads that are 10-100kb in size, while ONT can produce reads that are up to a megabase in size[5]. Both technologies have been used in the past to identify complex structural variants, construct genome assembly, and identify variants in difficult-to-map regions. These reads are long enough to cover the repeat regions and map better than the reads from NGS for those repeats that are larger than 300bps[5]. The read lengths also enable accurate examination of repeat numbers as well as sequences surrounding the repeat regions. Moreover, long-read sequencing allows direct examination of reads in the repeat regions and facilitates examining motif changes or repeat interruptions that are involved in novel STR diseases larger than the typical NGS read size (150-300bp). Some examples include the new pentanucleotide STRs found in SAMD12 that causes Benign Adult Familial Myoclonus Epilepsy (BAFME), and CAG repeat expansions without CAA interruptions that cause SCA4 in the *ZFHX3* gene[6, 7].

Another advantage of long-read sequencing is haplotype phasing. Reconstruction of haplotypes with NGS typically requires parental information, as the read length is often insufficient to cover multiple variants. Haplotypes can be inferred using statistical methods to bridge overlapping reads together, but in most cases, the blocks only span a few hundred bases around the targeted region due to limited read length[8]. Long-read sequencing allows construction of haplotype blocks without parental information, as the reads are long enough to span multiple variants and there is one variant per kb in the human genome on average. Haplotype blocks built from long-read sequencing spans from megabase to entire chromosomes and allow examination of long-range haplotypes around the regions[9, 10]. These long-distance haplotype blocks can phase repeat interruptions and repeat number information together with SNVs, allowing the quantification of minor allele frequency (MAF) based on repeat numbers or repeat interruptions. Recent studies show that haplotypes around the STR regions can be used for gene therapy targets, particularly for allele-specific gene editing that only edits the targeted allele since repeat regions are difficult to target without affecting the non-expanded allele. For example, a SNV in a patient cell line was used to specifically target expanded *C9orf72*, and the expanded HTT allele was specifically excised using CRISPR in vitro and in vivo, demonstrating the need for haplotype maps around the STR regions to facilitate sgRNA design[11–13].

ATXN2 is a protein with multiple roles, including, but not limited to, RNA metabolism, stress granule formation, and endocytosis. ATXN2 is expressed highly in the nervous system and has been found to associate with many neurologically- implicated proteins such as TDP-43, Parkin, and PABP. Previous studies also have implied that ATXN2 is necessary for proper circadian locomotor behavior and fatty acid pathways[14, 15]. ATXN2 contains CAG repeats in exon 1 that produce polyQ tracts when translated[16]. This CAG repeat region is associated with multiple diseases, including SCA2 and ALS and therapeutic interventions targeting *ATXN2* are underway[17, 18]. Fully-expanded repeats (polyQ > 35) without any CAA interruptions cause SCA2, and intermediate expansions (polyQ 27-33) are associated with ALS[19–21]. CAA interruption delays the age of onset in cases of Huntington’s Disease, and both this and its ability to cause entirely different diseases in *ATXN2* suggests its important role in the STR diseases[22]. Analyzing haplotypes around the *ATXN2* region is crucial, as accurately phasing the expanded repeats and associated interruptions with nearby SNVs enables precise identification of disease-related alleles. Such analyses provide deeper insights into phenotypic variability, differences in disease onset, and varying susceptibility among individuals affected by repeat-expansion diseases. Moreover, robust haplotype information is important for therapeutic strategies like allele-specific gene editing, where distinguishing pathogenic from normal alleles relies on SNV markers linked to the repeat expansions.

There have been a few haplotypes found surrounding *ATXN2*, in both non-disease and disease populations. Haplotype analysis in a South American population with SCA2 identified haplotypes around the regions, while analysis of Indian and Cuban populations identified haplotypes at two SNVs (G-T), rs695871 (C>G) and rs695872 (C>T) in both populations.[23–25] Analysis in a European population with ALS with intermediate expanded ATXN2 (27-33) also identified SNVs that formed haplotypes based on interruption numbers. Individuals with ALS who have one CAA interruption harbored C-C alleles, while individuals with ALS who have two or three CAA interruptions harbored G-T alleles[26]. However, these studies were done by statistical inference or by examining the SNVs that are close to known SNVs around repeat regions. Also, many of these studies were limited to specific populations such as European or Indian populations. In this study, we have discovered new haplotypes based on the interruption numbers in diverse populations using long-read sequencing datasets, extended the existing haplotypes from European populations, and examined the existing haplotypes that were previously reported in this region. Finally, we have discovered potential haplotypes that are specifically found in individuals with ALS who have three CAA interruptions.

## Methods

### Long-read datasets

Long-read datasets used in this study are from 1000G-ONT Sequencing Consortium and 1KG-Vienna Sequencing Consortium. 1000G-ONT Sequencing Consortium data were downloaded as bam files, while the 1KG-Vienna Sequencing Consortium data were downloaded as cram files. There were 100 bam files available in 1000G-ONT Sequencing Consortium at the time of download, and 1019 alignment files available for 1KG-Vienna Sequencing Consortium. Ancestral information was derived from 1000 Genome Project website, and the samples that were not found in the 1000 Genome Project website was directly searched in Coriell website. Samples with not enough coverage (min 4x) were filtered out as we could not form consensus sequences to identify repeat and interruption numbers or identify variants. There were total of 272 Americans, 294 Europeans, 303 East Asians, 336 South Asians and 521 Africans alleles in the study after filtering. In NYGC ALS samples, there were total of 4925 individuals but only 3846 individuals had *ATXN2* repeat size available. Out of 3846 individuals, 159 individuals had expanded *ATXN2* alleles (polyQ >= 27). After filtering for reads with expanded CAG repeats that are aligned into *ATXN2* regions, there were total of 154 individuals with expanded *ATXN2* alleles.

### NYGC ALS Consortium Cohort

For the phenotype examination, the metadata noted from NYGC ALS Sequencing Consortium was used, which used ExpansionHunter to estimate the repeat sizes and prior study has shown a correlation of 0.98 between EH and gold-standard repeat-primed PCR approaches for short repeats. Age at symptom onset was used to determine the age of onset. Ancestries were determined using columns that includes ancestry SNVs percentages, and most individuals had > 90% European ancestries. We also examined individuals with expanded *ATXN2* alleles to see whether they have other ALS causal mutations, and there were two individuals with SOD1 or C9orf72 mutations. Other than these two, other individuals did not contain any notable mutations.

### Penn Integrated Neurodegenerative Disease Database Cohort

The Penn INDD includes deep phenotyping and genomic characterization of thousands of individuals with neurodegenerative disease including, ALS, FTD, PD, and AD, as described in detail elsewhere. A subset of these individuals have been previously screened for *ATXN2* intermediate expansions using previously described methods ^12^ . Briefly we used a modified PCR cycling condition consisting of 2 min at 94 °C followed by cycles of 94 °C for 30 sec, annealing starting at 60°C for 30 sec, and extension at 72°C for 1 min, ending with a final extension step of 5 min at 72°C. The annealing temperature was decreased in 0.5°C steps for 14 cycles then at 53°C for 30 cycles. PCR products were separated by capillary electrophoresis with 23 sec injection time using POP-7 polymer and a 36 cm 16-capillary array on an ABI PRISM 3130xl Genetic Analyzer (ThermoFisher). Fragment sizes were determined by size standard Genescan ROX-500 with GeneMapper software v3.7.[27]

### Targeted Long-Read Sequencing in Penn INDD Cohort with Intermediate Expanded ATXN2

To amplify the number of read counts in the patient samples, long amplification PCR targeting around CAG regions of *ATXN2* with NEB’s LongAmp Hot Start Taq 2X Master Mix was used on 41 neurological diseases individuals with intermediate expanded *ATXN2* alleles. PolyQ sizes of the individuals varied from 27 – 33. For the forward primer sequence, GTTAGCCGGGACAACACTGA was used and for the reverse primer sequence, TGGGGGAAAGGAGAGCATTC was used to target 7077 bp region in chr12:111597669 - 111604746 that includes both three CAA ALS SNV at chr12:111603681 and the SNVs we found in the populations. PCR was performed using LongAmp Hot Start Taq 2X Master Mix (NEB, M0533) in 50 µL reactions containing 25 µL 2× master mix (1× final), 2 µL each of 10 µM forward and reverse primers (0.4 µM final each), 1 µL template DNA (∼250 ng), 1.5 µL DMSO (3% v/v), and nuclease-free water to volume (18.5 µL). Thermocycling conditions were: 94 °C for 30 s; 35 cycles of 94 °C for 15 s, 58 °C for 30 s, and 65 °C for 8 min; followed by a final extension at 65 °C for 10 min and a 4 °C hold. Amplified products were purified with AMPure XP beads (A63881) at a 0.4× beads-to-sample ratio, washed twice with 80% ethanol, air-dried, and eluted in 15 µL nuclease-free water. Following targeted PCR, 650ng of the PCR product was used for the standard protocol of Nanopore’s Native Barcoding Kit 24 V14 (SQK-NBD114.24). 48 samples, including 7 control samples and 41 patient samples, were pooled into two 24 sample libraries and sequenced with Flongles. After the alignment, the reads less than 5kb were filtered for accurate phasing between *ATXN2* CAG repeat region and SNVs. 8 samples were filtered out due to poor alignments or lack of SNVs and indels for phasing.

### SNV Calling and Phasing

Nanocaller(v3.4.0) was used to call SNVs and phase the variants around the *ATXN2* gene region. Nanocaller is a SNV caller for long-reads that utilizes deep convolutional neural network to detect SNVs/indels, builds long-range haplotype structure to predict each SNVs and phase variants with WhatsHap[28]. Nanocaller produced VCF files with phased variants information. Parameter used was –cpu 4 – preset ont –regions chr12:111098951-112099019 –mode snps --phase. Samples with not enough coverage were filtered out as they did not call variants in the region, and we were left with 1726 alleles in the end. In NYGC ALS datasets, HaplotypeCaller(v 3.5.0) was used to call SNVs. These SNVs were not phased to interruption numbers/polyQ numbers due to lack of read lengths in NGS.

### Identifying Repeat Interruption and Repeat Numbers

Consensus sequence was generated from alignment files using MUSCLE[29] and custom python script. Using the consensus sequence, the CAA interruption numbers, and CAG repeat numbers were directly counted using simple counting in *ATXN2* CAG repeat regions. For NYGC dataset, polyQ numbers from the metadata was used as the standard. The polyQ number was confirmed with ExpansionHunter as well[30]. Because the metadata counts CAG and CAA as same numbers, the aligned reads in *ATXN2* CAG repeat regions from the bam files were used to directly determine CAG repeat and CAA interruption numbers. For example, if there are 27 polyQ determined from the metadata and there are 24 CAG and 3 CAA in the reads, we determined that there were 24 CAGs and 3 CAAs in the allele with manual inspection.

### Identifying Top SNVs

We examined SNVs around the *ATXN2* regions, and calculated MAFs in population with one, two and three CAA interruptions. We calculated differences in allele frequencies for each SNVs and identified the top 10 alternate alleles with the biggest allele frequency differences between population with one CAA and two CAA interruptions. Three CAA interruptions were not included in the further population analysis as the number of alleles with three CAAs were too low.

For ALS samples, we calculated MAF in NYGC ALS dataset based on the CAA interruption numbers in the expanded allele and compared their allele frequencies with their counterpart for one CAA interruptions and two CAA interruptions in European populations in long-read datasets. We used the entire European population frequency for three CAA interruptions in the expanded allele in *ATXN2* individuals since there were not enough European with three CAA interruptions. We did not include 4 or 5 CAA interruption alleles as the number of samples were too low.

### Linkage Disequilibrium and Fisher’s Exact Test

Linkage disequilibrium was calculated using following the equation, where AF = allele frequency. r^2^ = [(AF_AB_ – (AF_A_* AF_B_))^2^ / (AF_A_ * (1-AF_A_) * AF_B_ * (1-AF_B_)). Fisher’s exact test and odds ratios were calculated using scipy.stat.fisher_exact function, with haplotype frequency as the one used for the calculation. Two-way Anova was calculated using sm.stats.anova.lm after separating allele frequency based on different interruption number and ethnicity.

### In silico functional genomic annotation of ATXN2-region variants

To evaluate whether *ATXN2*-region variants were associated with regulatory features relevant to ALS, we performed *in silico* annotation of rs148019457 and rs4098854 using publicly available QTL, chromatin interaction, TAD datasets harmonized using hipFG in FILER[31, 32]. Variant coordinates were analyzed in hg38, and annotations were restricted to records overlapping the variant-containing locus or chromatin/TAD intervals containing the variant. FILER metadata were used to map each annotation record to its original data source, assay type, tissue or cell type, and reported statistical metric.

For QTL annotation, we queried public eQTL and sQTL tracks for records implicating *ATXN2*. For each variant, we retained *ATXN2*-associated records and extracted the QTL type, data source, tissue or cell type, nominal p-value, and corrected statistic when available. GTEx v8 brain QTL records were interpreted as nominal associations because the queried GTEx tracks represented nominally significant eQTL/sQTL association files and did not provide FDR- or q-value–corrected statistics for the extracted ATXN2 records[33]. MiGA microglial sQTL records were interpreted using both nominal p-values and reported FDR values[34].

For chromatin interaction annotation, we identified chromatin loops in which the variant-containing anchor interacted with an anchor annotated to ATXN2 or the surrounding 12q24 regulatory neighborhood. For each interaction, we recorded the FILER track identifier, source dataset, assay, tissue or cell type, interacting anchor annotation, available confidence score or interaction score, and genomic distance when inferable from the interaction coordinates. The chromatin interaction datasets included promoter-capture Hi-C data from human DLPFC from Jung et al. [35], IM-PET interactions from middle hippocampus from 4DGenome[36], and neuronal PLAC-seq interactions from Nott et al [37]. Where available, quantitative confidence metrics were retained; for example, the IM-PET dataset reported confidence scores on a 0-1 scale. When p-values or FDR values were not present in the extracted annotation records, no corrected interaction significance was inferred.

For TAD annotation, we intersected rs148019457 and rs4098854 with TAD intervals from 3D Genome Browser annotations [38], including DLPFC and hippocampus datasets. For each TAD overlap, we recorded the TAD boundaries, whether *ATXN2* and neighboring genes fell within the same TAD, and the distance of each variant from the left and right TAD boundaries. The two variants were then compared to determine whether they occupied the same higher-order chromatin domain as *ATXN2* and whether chromatin interaction evidence occurred within that shared domain. All regulatory annotations were interpreted as gene-prioritization evidence rather than direct proof of variant function. QTL associations were considered statistically supported only when corrected statistics were available and met multiple-testing significance thresholds. Chromatin interaction and TAD annotations were interpreted as evidence of physical proximity or shared chromatin domain structure, but not as evidence that either variant directly alters *ATXN2* expression, splicing, or protein abundance.

## Results

### Summary of Datasets Used for *ATXN2* Haplotype and Repeat Analysis: 1000G Population, NYGC-ALS, and Penn Integrated Neurodegenerative Disease Database

We first obtained two long-read sequencing datasets from two different consortia, the 1000G-ONT Sequencing Consortium[39] (n = 100) and 1KG-ONT-Vienna Sequencing[40] Consortium (n = 1019). Both consortia examined samples from the 1000 Genomes Project, and contain diverse populations that include participants of European, East Asian, African, American and South Asian descent. The 1000G-ONT dataset was sequenced using the R9.4.1 flow cell with an average depth of 37.4x and a read N50 of 53.8kbp, and the 1KG-ONT Vienna dataset was sequenced with the R9.4.1 flow cell, with median coverage of 16.9x and N50 of 20.3kb. To build a consensus sequence and identify SNVs for phasing long-range haplotypes, some of the low coverage samples were filtered out (minimum 4x coverage across *ATXN2* region), and we were left with a total of 1726 alleles from 863 individuals[41]. After filtering, we had a total of 272 American, 294 European, 303 East Asian, 336 South Asian and 521 African alleles (Figure 1A).

**Figure 1.**
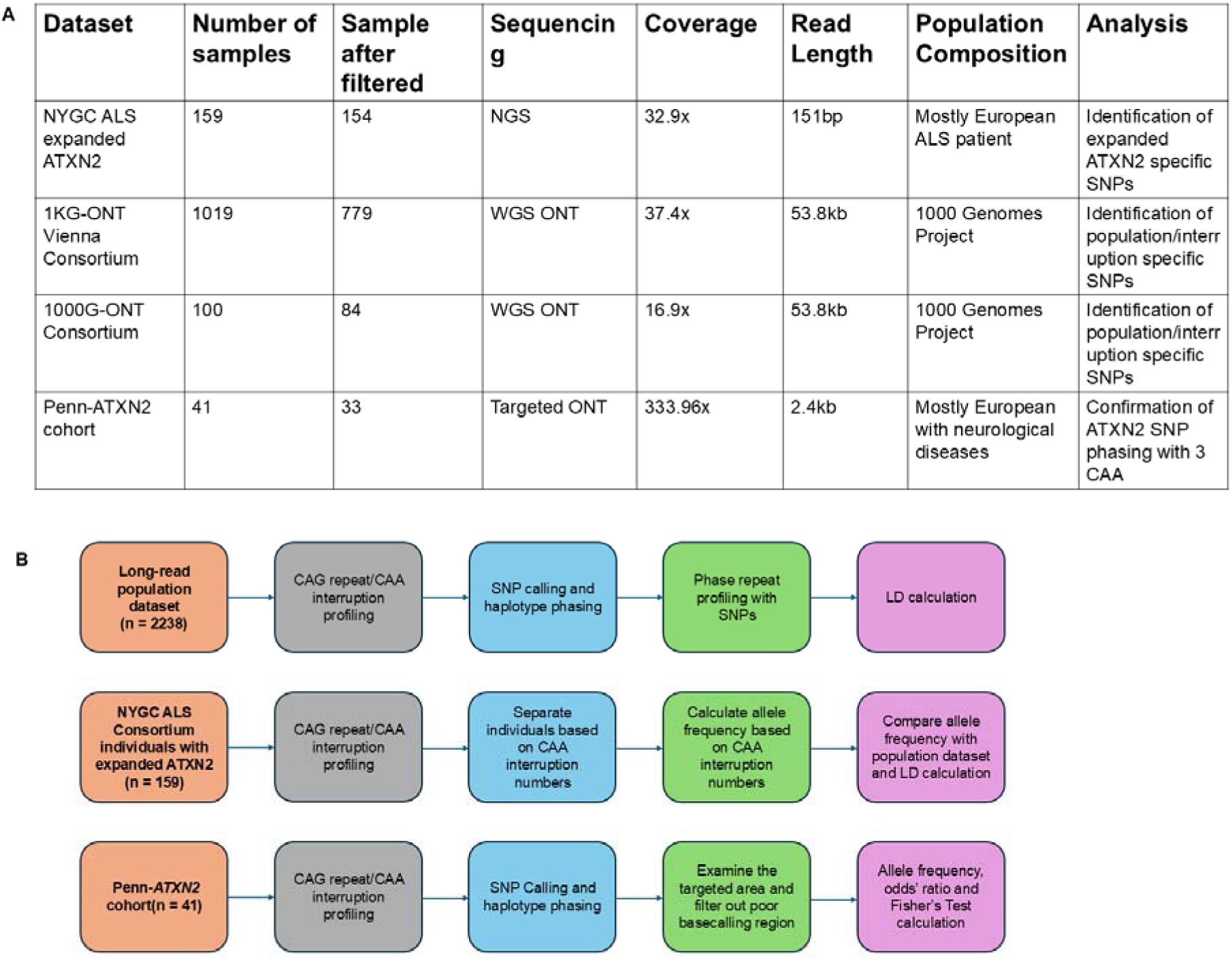
A) Table Showing Overview of Datasets. B) Workflow of Datasets.

The ALS Consortium dataset generated by the New York Genome Center (NYGC) was sequenced using HiSeq X or NovaSeq V1 and included a total of 4925 individuals with ALS Spectrum Motor Neuron Disease (MND), ALS Spectrum MND with other neurological disorders, other neurological disorders, and non-neurological controls. While this is a short-read dataset, the read length is sufficient to cover repeat regions completely for our inference of repeats. There was a total of 159 individuals with expanded *ATXN2* (CAG + CAA >= 27), with some individuals having expanded *ATXN2* repeats in both alleles. After filtering for sufficient coverage in *ATXN2* repeat regions, we were left with 154 individuals with expanded alleles in *ATXN2*. Most of the individuals (∼90%) with expanded *ATXN2* alleles were of European ancestry (Figure 1A).

We identified 41 individuals in the Penn Integrated Neurodegenerative Disease Database (INDD) with an intermediate expanded *ATXN2* repeat (27-33) from previous short-read screening. The observed phenotypes included ALS (N=21), Parkinson’s disease (N=10), dementia (N=4), frontotemporal degeneration (N=2) and other neurodegenerative diseases (N=4) (Table S1). We performed targeted ONT sequencing on *ATXN2* on these subjects. The median N50 was 2.4kb and the average read coverage around *ATXN2* regions was 2537bp. All but two individuals were of European ancestry. Eight samples were removed from the analysis due to inadequate read numbers or poor read quality in the *ATXN2* region, and therefore we focus our analyses on 33 individuals with neurological diseases meeting data quality standards (Figure 1A).

### *ATXN2* polyQ repeats and CAA Interruptions in 1000G and NYGC-ALS Datasets

We first examined the number of CAA interruptions within the *ATXN2* CAG+CAA repeats (that is, polyQ repeats) in the long-read 1000G population datasets. Traditional methods often refer to the number of glutamines (polyQ) in the *ATXN2* CAG repeat regions for diagnosis, because many studies use PCR-based methods (such as repeat-primed PCR and Fluorescent PCR Sizing) to quantify the repeat numbers without knowing the exact sequence. The exact number of CAA interruptions and CAG repeat expansion have not previously been characterized in diverse populations. We therefore decided to quantify the total polyQ (the sum of CAG repeat numbers and CAA interruption numbers), the CAG repeats, and the CAA interruptions in the *ATXN2* CAG region separately in order to examine the distribution of CAA interruption numbers and CAG repeat numbers in the 1000G population and the ALS cohort.

In the 1000G dataset, the most common number of polyQ is 22, followed by 23, which is similar to the results shown in previous studies in *ATXN2* CAG region[42, 43]. African populations had a higher percentage of non-22 polyQ alleles compared to the other populations (Figure S3A). There were significant differences between the number of polyQs in populations with one CAA interruption vs two CAA interruptions, and those with three CAA interruptions had expanded polyQ compared to the population with one or two CAA interruptions (Figure 2A). The most common number of CAA interruptions was 2 (66%), followed by 1 (33%). Three CAA interruptions were present at a low frequency (< 1%) in the population. East Asians had the highest percentage of alleles with 1 CAA interruption, while South Asians had the highest percentage of alleles with 2 CAA interruptions (Figure S3C,E). East Asians had no alleles with 3 CAA interruptions in this dataset, while the rest of the populations had 1-2% of alleles with 3 CAA interruptions. The East Asian population was significantly different from European, African, and South Asian populations, but not from the American population. No other significant differences were found among the groups. The most common number of CAG repeats was 20, followed by 21, as the most common number of polyQ was 22, which harbored either 1 or 2 CAA interruptions (Figure 2A). As mentioned previously, the distributions were similar to that of polyQs.

**Figure 2.**
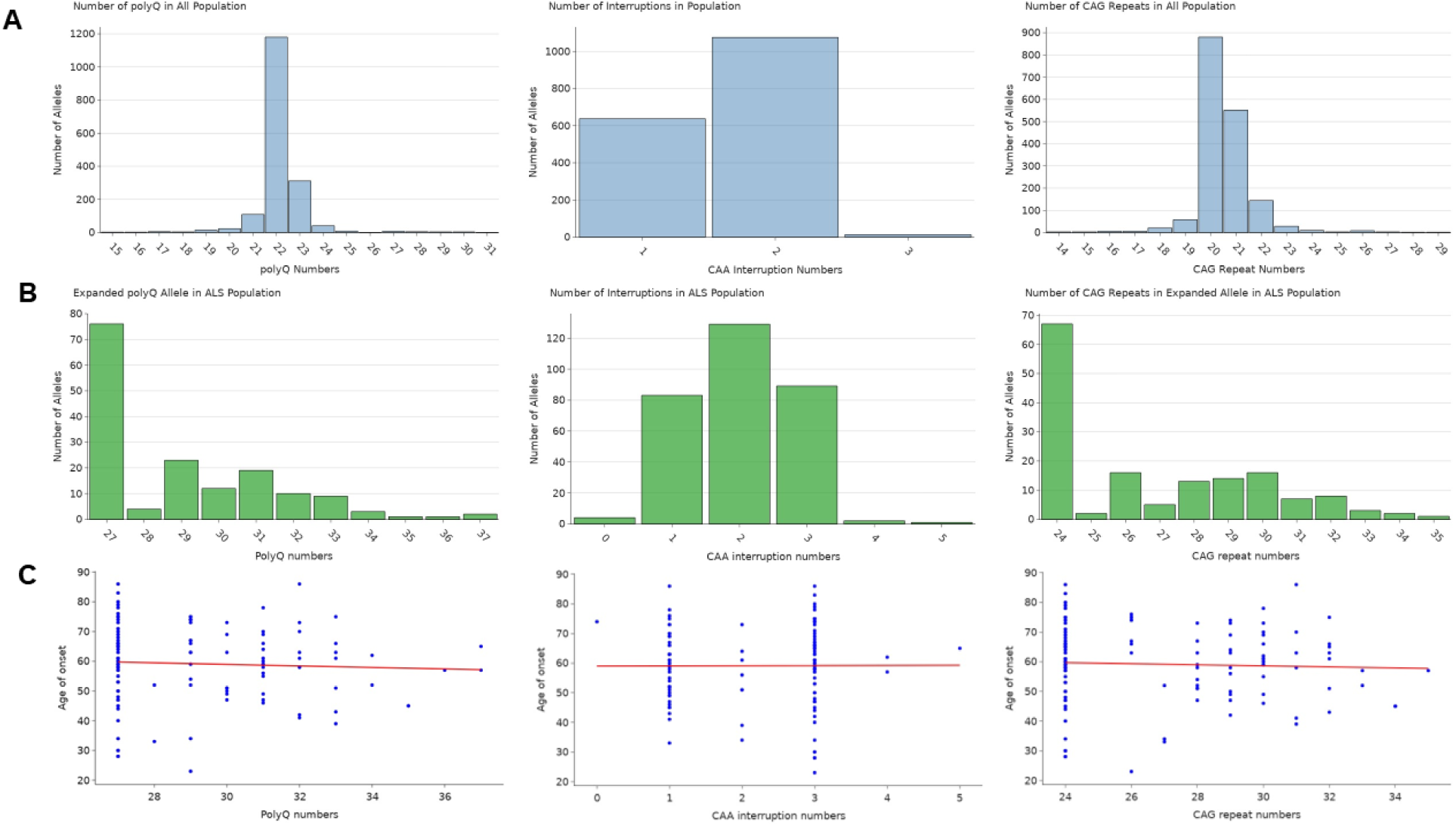
Overview of Number of PolyQ, CAA interruptions and CAG repeats in *ATXN2* CAG repeat regions A) Distribution of number of PolyQ, CAA interruptions and CAG repeats in the Long-Read Population dataset. B) Distribution of number of polyQ, CAA interruptions and CAG repeats in expanded *ATXN2* allele of NYGC ALS population with expanded *ATXN2*. C) Scatter plots of age of onset (Y-axis) In relationship with number of interruptions, CAG repeats and polyQ (X-axis). Red line shows linear regression of age of onset with X-axis.

In the expanded alleles of NYGC-ALS individuals with intermediate *ATXN2* alleles, the most common number of polyQ was 27 (N=76), followed by 29 (N=23). The highest number of polyQ was 37, only observed in 2 individuals. As seen in previous studies, the frequency of polyQ > 27 was significantly higher in NYGC-ALS individuals compared to 1000G population datasets (P-value = 0.0011), with odds ratios of 2.00 (95% CI, 1.28 – 3.28). Surprisingly, the most common number of CAA interruptions was 3 (N=84, 54.5%), followed by 1 CAA interruption (N=51, 33.1%). 2 CAA interruptions were present at a much lower frequency (N=13, 8.4%), and there were rare 4 or 5 CAA interruptions (N=3, 1.9%) that were only observed in neurological disease but not neurological control individuals. The number of interruptions was significantly higher (p-value < 1e-10, t-test) in the *ATXN2* expanded population with ALS compared to the control population, whose interruption numbers were mostly 1 or 2. The most common number of CAG repeats was 24 (N=67, 43.5%), followed by 26 and 30 (N=16, 10.4%), as most of the expanded *ATXN2* alleles with 27 polyQ had 3 CAA interruptions (N=67, 43.5%) (Figure 2B). Because previous studies have reported that the number of CAA interruptions are positively correlated with age of onset in ALS and the number of CAG repeats correlates with age of onset in Huntington’s disease[22, 26], we examined the relationship between the number of CAG repeats and CAA interruptions with age of onset. There was no significant relationship between the number of CAA interruptions and age of onset, and there is a slight but non-significant inverse correlation between the number of CAG repeats and the age of onset (r = -0.043) (Figure 2C). Overall, the ALS population had a higher number of CAA interruptions than the control population, but we did not replicate a significant correlation between CAA interruptions and the age of onset that was previously suggested.

### Interruption-Associated SNVs and Haplotype Block Structures

To assess whether CAA interruption number is associated with distinct haplotype backgrounds, we analyzed single nucleotide variants (SNVs) within 1 Mb of the *ATXN2* locus and identified those whose allele frequencies differed according to interruption count in long-read population datasets. We identified ten SNVs that were enriched in alleles with two CAA interruptions compared to the reference (one CAA). Alleles with three CAA interruptions shared a nearly identical pattern to those with two interruptions. Five SNVs showed consistent frequencies across diverse populations, while the other five SNVs showed variations in frequency depending on the ethnicity (Figure S1). Linkage disequilibrium (LD) analysis revealed that these ten SNVs organize into two distinct haplotype blocks. The first block, C-C-C-G, is formed by rs10774629, rs10774631, rs7398196 and rs10849962, and the second block, T-G-T-G, is comprised of rs695871, rs695872, rs7953810 and rs12810456. These two haplotype blocks are merged in European, South Asian and American populations, but are broken into two sub-haplotype blocks in African and East Asian cohorts, suggesting haplotype structures are affected by interruption status and population history at the same time (Figure S). We also examined another haplotype around the *ATXN2* region based on the South American SCA2 population, and found that the prevalence of the common haplotype A-G-C-C-C (non-SCA2 haplotype) and G-C-G-A-T (SCA2 haplotype) in this study was low in our South American population (Table S2G).

We further examined both haplotype blocks based on their interruption numbers and ancestry. The T-G-T-G haplotype, which includes two SNVs (rs695871 and rs695872) that were previously found in Cuban and Indian populations with SCA2 and European populations with three CAA interruptions with ALS, were present with high frequency in European, South Asian and American populations, but was at much lower frequency in East Asian and African populations. Even at two CAA interruptions, the T-G-T-G haplotype was a minor haplotype compared to the reference haplotype C-C-C-A, showing that ethnicity plays a stronger role than the number of CAA interruptions in this haplotype block. In contrast, the C-C-C-G haplotype was presented at high frequency in populations with two CAA interruptions regardless of ethnicity (Table S2A,B,C,D,E,F). While the haplotype frequency was variable in the populations with one CAA interruption, all populations showed higher frequency of the C-C-C-G haplotype in two CAA interruptions, while the frequency of C-C-C-G haplotype was lower in one CAA interruption, suggesting that this haplotype block is affected by the number of CAA interruptions rather than ethnicity. Fisher’s exact test based on the haplotype frequency between the population confirmed the differences between the haplotype block. The haplotype frequency for C-C-C-G differed significantly when the interruption number was different (American with one CAA interruptions vs American with two CAA interruptions p-value =6e-33), but not based on the ethnicity (American with two CAA interruptions vs European with two CAA interruptions p-value =0.53). At the same number of interruptions, the difference between populations was insignificant for most populations (Figure 3A). The haplotype T-G-T-G had significant differences in frequency even at the same number of interruptions depending on the ethnicity (American with two CAA interruptions vs European with two CAA interruptions p-value =2.1e-11). (Figure 3B). Comparison of odds ratio showed similar results (Figure 3C,D), and two-way ANOVA tests confirmed that the frequency differences in C-C-C-G haplotype was due to CAA interruption number, while the differences in T-G-T-G haplotype was due to a mixture of CAA interruption number and ancestry.(Table S3A,B)

**Figure 3.**
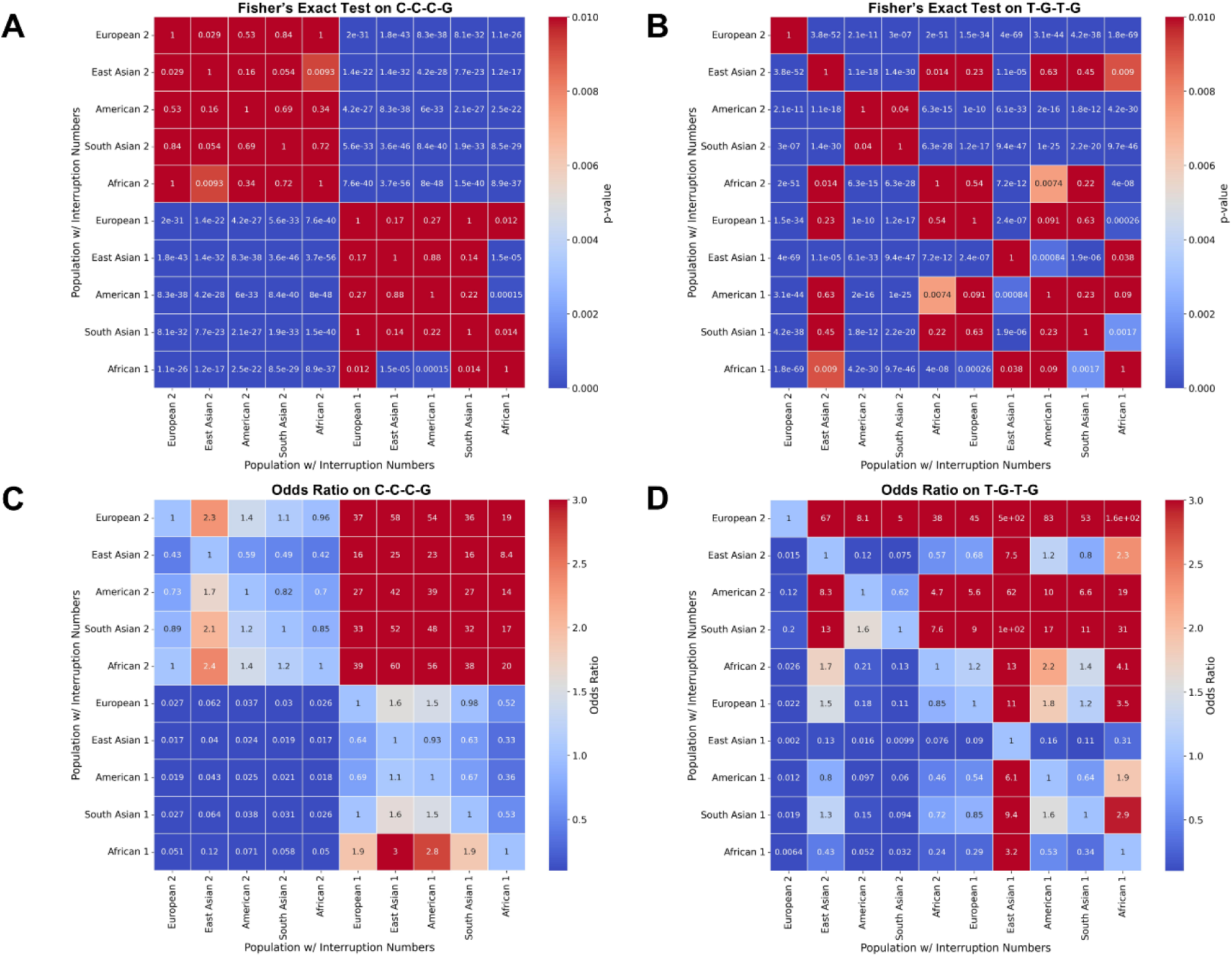
Heatmaps showing statistical tests result. A) Heatmap of Fisher’s exact test for haplotype C-C-C-G, in different ethnicity with one CAA interruption and two CAA interruption. B) Heatmap of Fisher’s exact test for haplotype T-G-T-G, in different ethnicity with one CAA interruption and two CAA interruptions. C) Heatmap of odds ratio on haplotype C-C-C-G, in different ethnicity with one CAA interruption and two CAA interruption. D) Heatmap of odds ratio on haplotype T-G-T-G, in different ethnicity with one CAA interruption and two CAA interruption

### ALS Specific SNVs and Haplotype Signatures in Expanded *ATXN2* Alleles

We first confirmed the frequency of haplotypes that we have identified in previous sections in the NYGC-ALS population with expanded *ATXN2* alleles. We first calculated the r^2^ between the top 10 SNVs, and the heatmap resembled that of the European LD correlation coefficient heatmap, with most SNVs in LD with each other. We further analyzed haplotypes T-G-T-G and C-C-C-G and examined their minor allele frequency. As expected from the European population, T-G-T-G was the major haplotype while C-C-C-G was also the major haplotype. We performed Fisher’s exact test to compare SNV frequencies between long-read population datasets and the NYGC-ALS dataset, and found that there was not a significant difference between the two datasets in these top 10 SNVs (Figure 4A).

**Figure 4.**
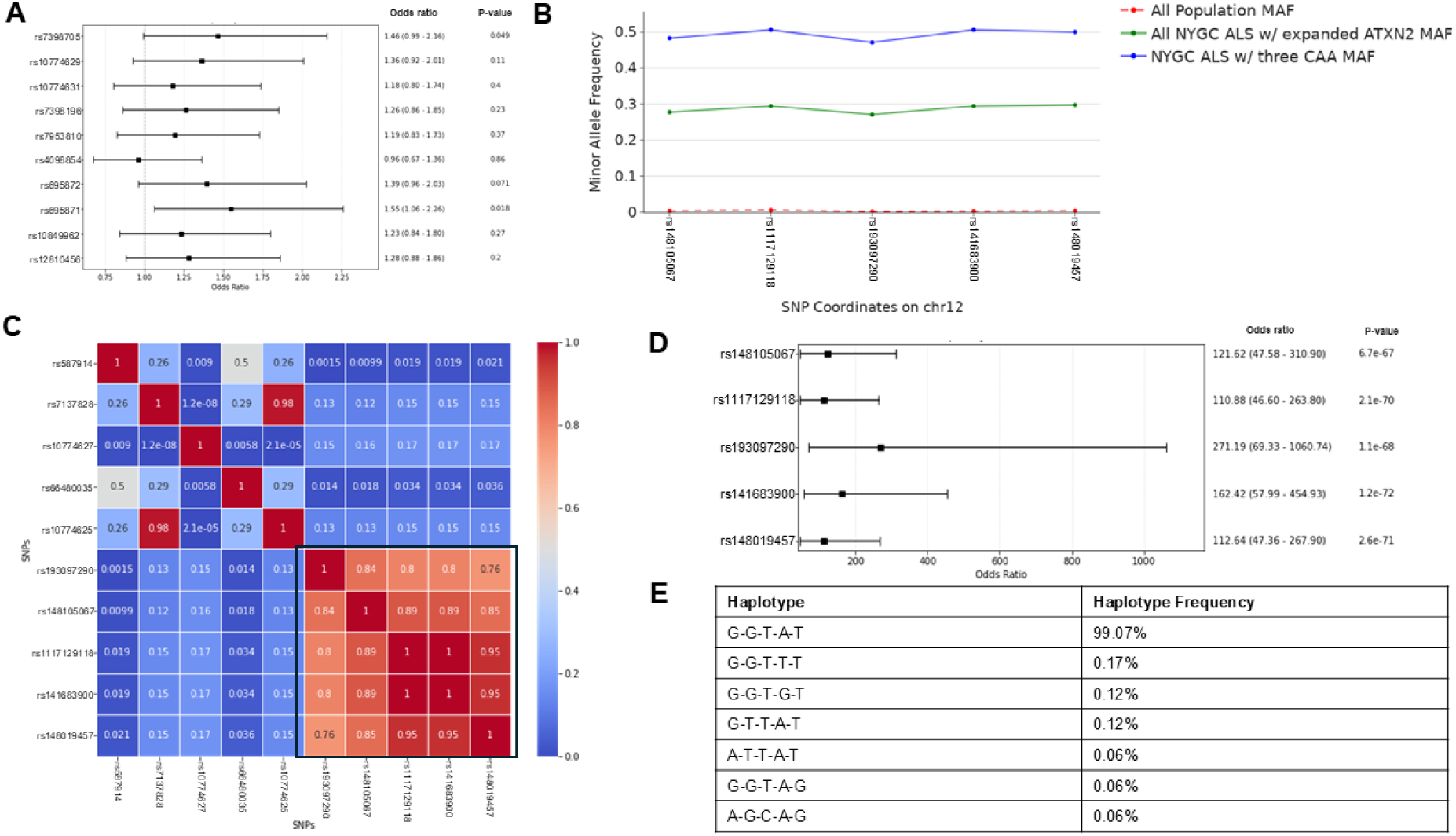
Overview of haplotype and alternate allele in NYGC ALS population. A) Forest plot showing odds ratio and Fisher’s test results of top 10 SNVs frequencies between the NYGC ALS patient datasets and long-read population datasets. B) Allele frequency of top SNVs found ALS patients for long-read datasets, all NYGC ALS patients with expanded *ATXN2* alleles and all NYGC ALS patients with three CAA interruptions. C) Heatmap of LD correlation coefficient of top alternate allele more commonly found in NYGC ALS population compared to long-read population. D) Forest plot showing odds ratio and Fisher’s test results of top ALS SNVs frequencies between the NYGC ALS patient datasets and long-read population datasets. E) Haplotype frequency in long- read dataset population.

We examined some of the top alternative alleles that were found in the NYGC-ALS population compared to our long-read population datasets using allele count. While it is not possible to phase the different interruption numbers into different alleles due to limitations of short reads, we used simple allele count differences. There were 5 alternate alleles at rs148105067, rs117129118, rs193097290, rs141683900 and rs148019457 that were found to have the greatest difference between ALS and non-ALS populations (Figure 4B,C, Table 1). These alternate alleles are rare alleles that are found in only 1% or less in the population. We also examined the MAF in the gnomAD database[44], and found that the European population, especially Finnish, had higher MAF than the non-European population (Table 1). Further analysis showed that these alternate alleles are mostly found in the patients that have three CAA interruptions. Interestingly, these alleles were present in almost all the patients with three CAA interruptions. Linkage disequilibrium analysis showed that these 5 alleles were found to be highly correlated with each other as well (Figure 4C,D). Both odds ratios test and Fisher’s exact test showed that their MAF differed significantly from each other, with odds ratio > 100 and p-value < 1e-10 for all SNVs. In the long-read dataset, these alternate alleles were only present in European, American and African populations. Interestingly, these alternate alleles were present at a much higher frequency in the population with three CAA interruptions (N=14, 1.6%) relative to population without three CAA interruptions in the long-read dataset, but the haplotype frequency for C-C-G-A-T, which is made up of the alternate alleles, was very low, as it was only found once in the long-read dataset population. The most found haplotype was A-T-A-C-C, the haplotype made up of reference alleles, which made up 99% of the haplotype in the population (Figure 4E). Only individual in the control population with C-C-G-A-T haplotype was found in European with three CAA interruptions, who has polyQ number of 28 (25 CAG repeats and 3 CAA interruptions). Overall, we have discovered SNVs and potential haplotypes that may be present specifically in individuals with ALS that have three CAA interruptions in European populations.

**Table 1.** Allele frequency of top 5 SNVs found in ALS patients with three CAA interruptions in expanded *ATXN2* allele.

| SNV | AF, all<br>ALS<br>Patient<br>s | AF, non-three CAA<br>interruptio<br>n ALS<br>patients | AF, three<br>CAA<br>interruption<br>s ALS<br>patients | AF,<br>gnomAD<br>all<br>populatio<br>n | AF,<br>gnomAD<br>non-Finnish<br>European<br>Populatio<br>n | AF,<br>gnomAD<br>Finnish<br>populatio<br>n |
| --- | --- | --- | --- | --- | --- | --- |
| rs148105067 | 0.278 | 0.015 | 0.483 | 0.00685 | 0.01401 | 0.04170 |
| rs117129118 | 0.294 | 0.022 | 0.506 | 0.0103 | 0.01718 | 0.04222 |
| rs193097290 | 0.271 | 0.015 | 0.471 | 0.0342 | 0.01535 | 0.04450 |
| rs141683900 | 0.294 | 0.022 | 0.506 | 0.0103 | 0.01720 | 0.04255 |
| rs148019457 | 0.297 | 0.037 | 0.500 | 0.0103 | 0.01746 | 0.04409 |

### Targeted ONT Sequencing Confirms Presence of SNV in Intermediate *ATXN2* Alleles with 3 CAA Interruptions

To further confirm the presence of these SNVs in the ATXN2 expanded allele with three CAA interruptions, we performed targeted long-read sequencing for a group of 41 individuals affected with neurological diseases. The samples from these individuals were obtained from the Penn INDD cohort with intermediate expanded ATXN2 alleles from previous genetic testing (hereafter referred to as the “Penn-ATXN2” cohort) (Figure 5A). In the expanded *ATXN2* alleles, three CAA interruptions made up the majority of the alleles (82%), followed by one and two CAA interruptions(18%), showing that the three CAA interruption makes up the majority of alleles in the intermediate expanded *ATXN2*. The majority of patients were of European descent except for two patients, so we used European populations from long-read population datasets to compare with the neurological patient cohorts. Because PCR focused on the region between chr12:111597669 – 111604746, we examined the SNVs that were present in this region including rs128508, rs695871, rs10849962, rs12810456 and rs148019457.

**Figure 5.**
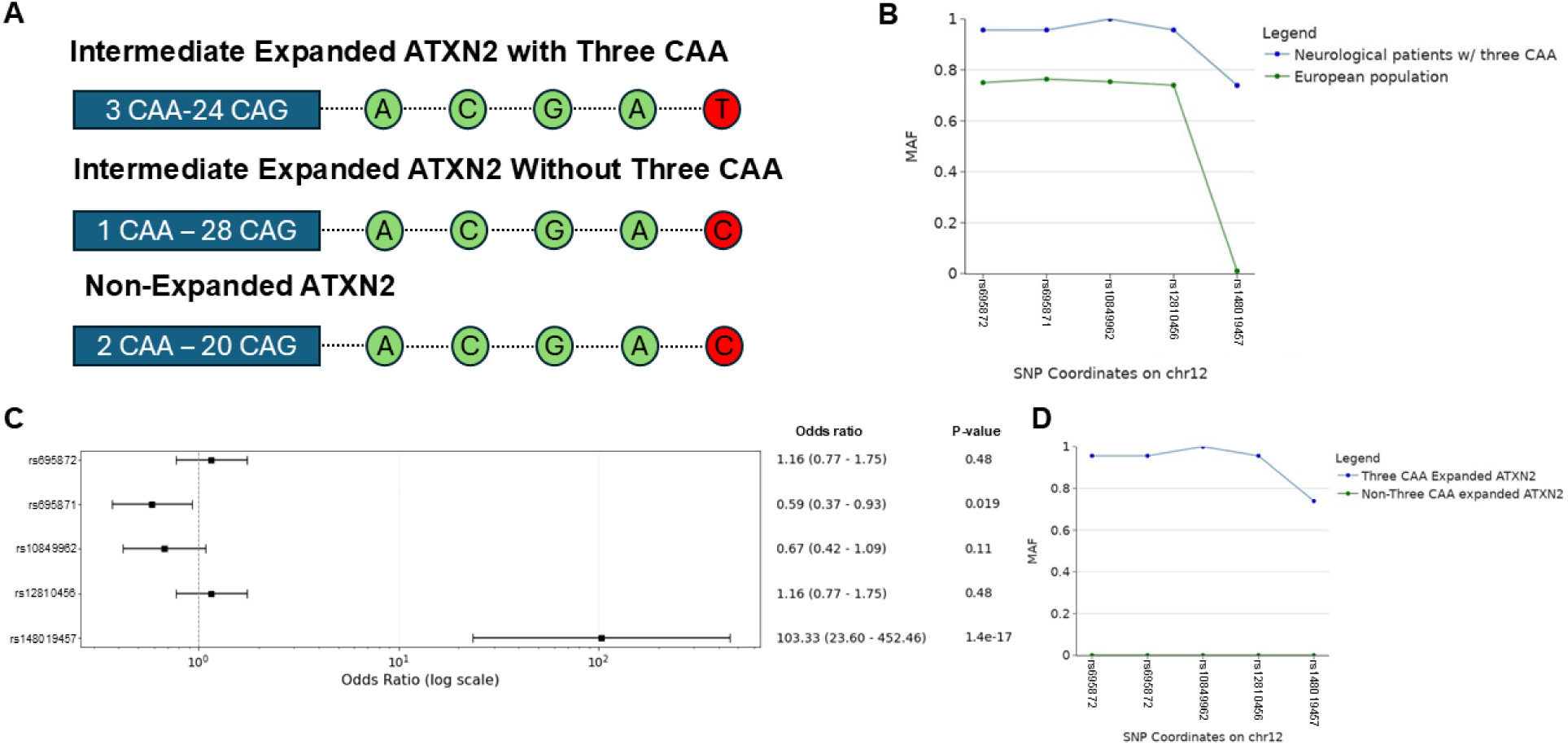
Targeted ONT sequencing in *ATXN2* CAG repeat regions to phase the expanded *ATXN2* CAG with three CAA interruptions and the SNVs for direct comparison. A) Expanded *ATXN2* CAG regions targeted sequencing schema. B) Allele frequency of the targeted SNVs in expanded *ATXN2* allele of neurological patients with three CAA interruptions and European population. AF_NEU_ represents Penn cohort allele frequency, while AF_EUR_ represent European population allele frequency. C) Forest plot showing odds ratio test and p-value for Fisher’s exact test between the expanded *ATXN2* allele of neurological patients with three CAA interruptions and European population in long-read dataset. D) Allele frequency of targeted SNVs in the expanded *ATXN2* allele of neurological patients with three CAA interruptions and the expanded *ATXN2* allele of patients without three CAA interruptions.

The MAFs for these SNVs in the expanded *ATXN2* alleles in neurological disease individuals with three CAA interruptions compared to neurological control individuals were similar for rs128508, rs695871, rs10849962 and rs12810456 but differed significantly between groups for rs148019457 (AF_NEU_ = 73.9%, AF_EUR_ = 1.0%) (Figure 5B). Odds ratio and Fisher’s exact test confirmed these, as none of the population SNVs (rs128508, rs695871, rs10849962 and rs12810456) were significantly different at a p-value set to 0.01, and odds ratios fell within the 0.5 – 1.2 range (Figure 5C). In contrast, the MAF for rs148019457 was significantly different (p-value = 1.4e-17) and had a greater odds ratio difference (103.33, CI 95% 23.60- 452.46) when comparing European population and Penn INDD cohort. As seen in the previous analysis, rs148019457 was not present in any of the expanded *ATXN2* alleles without three CAA interruptions, even in the neurological disease group, and was presented at a much higher rate in the expanded *ATXN2* allele with three CAA interruptions (Figure 5D). This SNV was uniquely found in neurological disease individuals with three CAA interruptions. Other population SNVs were also lacking in patients with non-three CAA interruptions, but this is due to the high presence of one CAA interruption allele in this particular cohort. Overall, we confirmed that rs148019457-G allele uniquely tags the expanded alleles with three interruptions in the Penn-ATXN2 dataset.

We have also examined the potential effects of our variants in gene expression and epigenetic change using publicly available datasets. We found chromatin interaction signals on our variants within an *ATXN2*-associated regulatory neighborhood. From a human dorsolateral prefrontal cortex (DLPFC) promoter capture Hi-C datasets, the rs148019457 containing region had an anchor annotated to *ATXN2* promoter, and more distal genes such as *SH2B3*, *BRAP*, and *ACAD10*[35]. An IM-PET dataset from middle hippocampus tissue and a neuronal PLAC-seq dataset placed the rs4098854- containing region in interactions involving *ATXN2*-annotated anchors and neighboring genes including *SH2B3*, *BRAP*, *ACAD10*, and *TRAFD*[36, 37]. IM-PET dataset showed high-confidence score (>0.85 and approaching 0.99, max = 1) for chromatin interaction involving *ATXN2*-associated interactions. Furthermore, topologically associating domain (TAD) analysis (data from the 3D Genome Browser, including DLPFC and hippocampus datasets) supports these evidences, as these variants fall in TAD around chr12:110.9Mb-112.4Mb that contains *ATXN2*, *SH2B3*, *BRAP*, *ACAD10*, and *MAPKAPK5*, showing that there are chromatin interaction changes in neuronal tissues in these regions[38]. Gene expression analysis showed limited evidences for direct involvement of our variants in regulation of *ATXN2*. rs148019457 was identified as a nominal *ATXN2* expression quantitative trait loci (eQTL) or splicing QTL (sQTL) across multiple GTEx v8 brain tissues, and rs4098854 showed nominal *ATXN2* sQTL associations in MiGA microglia from medial frontal gyrus[34]. However, the GTEx records did not include FDR- or q-value-corrected significance statistics for rs148019457 and rs4098854 showed high FDR values and were not significantly supported after multiple-testing correction.

## Discussion

Our study reveals potential haplotypes and SNVs around *ATXN2* based on different number of interruptions. Unlike short-read NGS, which provides limited phasing information between repeat regions and surrounding SNVs, long-read sequencing allows examination of haplotypes surrounding repeat regions based on different properties of repeat regions. We have extended the previously discovered haplotypes that were found in European, Cuban and Indian populations, and identified potential haplotypes based on two CAA interruptions in the CAG repeat region. Finally, we have discovered a potential new haplotype in ALS patients with expanded *ATXN2* in CAG repeat regions with three CAA interruptions. In this study, long-read sequencing has allowed us to examine full haplotypes and build haplotype blocks around the *ATXN2* region across diverse populations, allowing us to separate each haplotype based on different repeat interruption numbers and repeat numbers. We were able to distinguish the haplotypes based on different ethnicities as well, thanks to the diverse populations that were available in the 1000 Genomes Project. Furthermore, we were able to show that the previously published haplotypes discovered by the statistical inference methods in South American populations with no SCA2 were not present in our South American population due to a further SNP not phasing with other SNPs, showing the power of the long-read sequencing over the statistical inference methods with array analysis.

One particularly interesting point regarding the newly discovered haplotype (C-C-C-G) in populations with two CAA interruptions is how shared it is between ethnic populations based solely on interruption number. This suggests that this haplotype was founded at a much earlier time point, as it was found in all populations including African which usually has a diverse haplotype[45]. CAG repeat regions are prone to strand-slipping that results in repeat expansion, but this strand-slipping can be stabilized by CAA repeat interruption[46]. Furthermore, even numbers of CAG repeats are favored over odd numbers for a paired-end tetraloop that confers greater stability, as seen in the greater number of even-numbered CAG repeats in the population with two CAA interruptions. This could imply the importance of the haplotype, as two CAA interruptions are discovered at a higher rate, along with the haplotype C-C-C-G.

There are therapeutic implications of the haplotype as well. Allele specific gene editing using CRISPR-Cas9 often requires heterozygous SNVs around a protospacer adjacent motif (PAM) sequence following the sgRNA/DNA complementary region. In the ALS patients with expanded *ATXN2* CAG repeat regions, three CAA interruptions are much more frequent, and we have identified SNVs around the *ATXN2* gene region that are much more frequently found in patients with three CAA interruptions. These SNVs can potentially be used as a gene editing site to specifically remove the expanded allele, which has been shown in model organisms *in vivo* to effectively improve disease phenotypes[47, 48]. While there are limited evidence for gene expression changes, some of the SNPs are in the regions that are associated with chromatin interactions in brain tissues, suggesting that these variants may affect epigenetical impact. Interestingly, the five SNVs that we identified have higher frequencies in the Finnish population, and the Finnish population has a higher incidence of ALS compared to other world populations[49]. There have been no studies examining the frequency of intermediate *ATXN2* CAG repeat expansions in the Finnish population, but based on our analysis, there is a possibility that the Finnish population may have a higher percentage of expanded *ATXN2* with three CAA interruptions. Considering that these SNVs are specifically found to be more common in ALS populations, some Finnish ALS cases may be caused by expansion in *ATXN2*, and more tests to examine these regions are needed.

Furthermore, 4 of these SNVs (rs148105067, rs117129118, rs141683900 and rs193097290) are included in the Illumina Neuro Consortium and have previously been screened. One of the SNVs, rs117129118, is included in multiple SNV microarrays such as the Illumina Infinium Global Diversity Array. As previously mentioned, current analysis for *ATXN2* expansion identification requires targeted approaches or whole genome sequencing with bioinformatics experience. As seen in our analysis, these SNVs have a very high correlation with the intermediate *ATXN2* expansion in ALS patients with three CAA interruptions. Some of the SNVs could be used as a biomarker for potential future screening for ALS or patient selection for *ATXN2*-targeted clinical trials, especially for rs117129118 that is included in multiple common SNV array analysis as many patients would have been screened already. Considering the high presence of three CAA interruptions in intermediate *ATXN2* expanded patients, this biomarker may be applicable to hundreds of ALS patients that have been screened already or will be screened.

Our study comes with some limitations. While long-read sequencing allows direct phasing of repeats and surrounding SNVs, there are potential basecalling errors that could affect variant detection in some alleles. Furthermore, comparison of variants in different sequencing technologies may result in technical batch effects, as there may be different type of technological biases present in each cohorts. Our comparison of populations is also limited by biases, as the comparison between the general population and the NYGC-ALS population with high frequencies of ALS incidences may reflect genetic risk factors and ascertainment biases. NYGC-ALS population is enriched in population with specific clinical phenotypes, ALS, whereas the 1000 Genomes Project has not been screened for neurological condition. SNVs identified in expanded *ATXN2* alleles may also be in LD with unidentified ALS risk loci in NYGC-ALS population as well.

## Conclusions

In summary, we used long-read sequencing to simultaneously phase haplotypes, CAG repeat numbers, and CAA interruptions around *ATXN2* across diverse populations, allowing us to extend previously reported haplotypes and to resolve new ones defined by interruption number. We found that three CAA interruptions are rare in control populations across multiple ancestries but common among individuals with ALS who carry intermediate repeats, and that this interruption status can be tagged by an SNV in individuals of European ancestry and independently confirmed by targeted ONT resequencing. Because this tag SNV is captured on commonly used genotyping arrays, interruption status may be inferred from existing microarray data without targeted or long-read sequencing. Taken together, our findings show the value of interruption-aware, long-read haplotype phasing at repeat-expansion loci and its potential to support the development of precision genomic medicine for neurological disorders.

## List of Abbreviations

AFR: African (1000 Genomes superpopulation)
ALS: amyotrophic lateral sclerosis
BAFME: benign adult familial myoclonus epilepsy
Cas9: CRISPR-associated protein 9
CRISPR: clustered regularly interspaced short palindromic repeats
EAS: East Asian (1000 Genomes superpopulation)
EUR: European (1000 Genomes superpopulation)
FTD: frontotemporal dementia
HD: Huntington’s disease
INDD: Integrated Neurodegenerative Disease Database
LD: linkage disequilibrium
MAF: minor allele frequency
MND: motor neuron disease
NGS: next-generation sequencing
NYGC: New York Genome Center
ONT: Oxford Nanopore Technologies
PABP: poly(A)-binding protein
PacBio: Pacific Biosciences
PAM: protospacer adjacent motif
PCR: polymerase chain reaction
polyQ: polyglutamine
SAS: South Asian (1000 Genomes superpopulation)
SCA2: spinocerebellar ataxia type 2
sgRNA: single guide RNA
SMRT: single-molecule real-time
SNP: single nucleotide polymorphism
SNV: single nucleotide variant
STR: short tandem repeat
TDP-43: TAR DNA-binding protein 43
VCF: variant call format

## Consent for publications

Not applicable

## Authors’ contributions

K.W conceptualized and designed the study. J.C generated the Penn INDD ONT dataset. B.H.L and Y.Y.L conducted data analysis and interpreted the results. C.M, Y.S., D.A, and K.W. advised on the study and facilitated data annotation. K.W. supervised the study. B.H.L. drafted the initial manuscript. All authors read and approved the manuscript.

## Ethics approval and consent to participate

This study was approved by the Institutional Review Board of the Children’s Hospital of Philadelphia. All analyses were performed using de-identified data, and the requirement for informed consent was waived.

## Data Availability

All data produced in the present study are available upon reasonable request to the authors

## Acknowledgements

We are grateful to 1000G-ONT Sequencing Consortium and 1KG-ONT Vienna Sequencing Consortium for their public datasets. We thank the individuals affected with neurological diseases and ALS who have participated in research and contributed their DNA samples to research through NYGC ALS Sequencing Consortium and Penn Integrated Neurodegenerative Disease Database. We are also grateful to Eunran Suh and Vivianna Van Deerlin, and Molecular Integration in Neurological Diagnosis Initiative at University of Pennsylvania for providing DNA samples. All NYGC ALS Consortium activities are supported by the ALS Association (ALSA, 19-SI-459), the Tow Foundation, The Target ALS Human Postmortem Tissue Core and New York Genome Center for Genomics of Neurodegenerative Disease.

## Availability of data and materials

The scripts used for data analysis is available at https://github.com/WGLab/Project_atxn2_population_analysis. 1000G-ONT data is available at https://millerlaboratory.com/1KGP-LRS.html and 1KG-ONT Vienna data is available at https://www.internationalgenome.org/data-portal/data-collection/1kg_ont_vienna.

---

**Figure S1.**
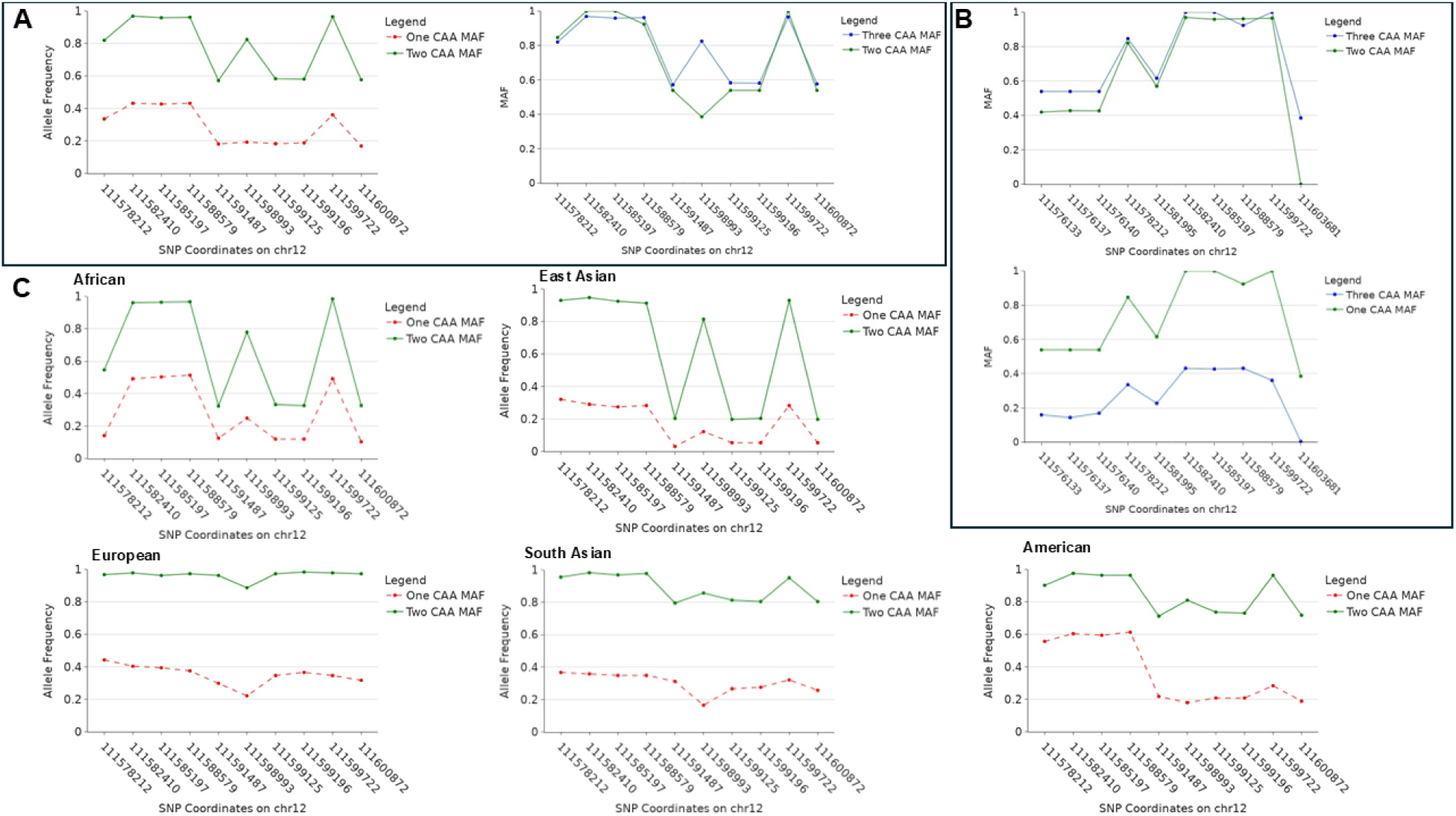
Overview of Top 10 MAFs in Long-Read Population Dataset. A) Top 10 minor alleles that are more commonly found in allele with two CAA Interruptions in *ATXN2* compared to alleles with one CAA interruption comparing their frequencies in population with one CAA interruption and two CAA interruptions (left) and their frequencies in population with two CAA interruptions and three CAA interruptions (right). B) Top 10 minor alleles that are more commonly found in alleles with three CAA interruptions in *ATXN2* compared to alleles with one CAA interruption and their frequencies in population with one CAA interruption (bottom) and their frequencies in population with two CAA interruptions (top). C) Top 10 MAFs in African, American, East Asian, South Asian and European population.

**Figure S2.**
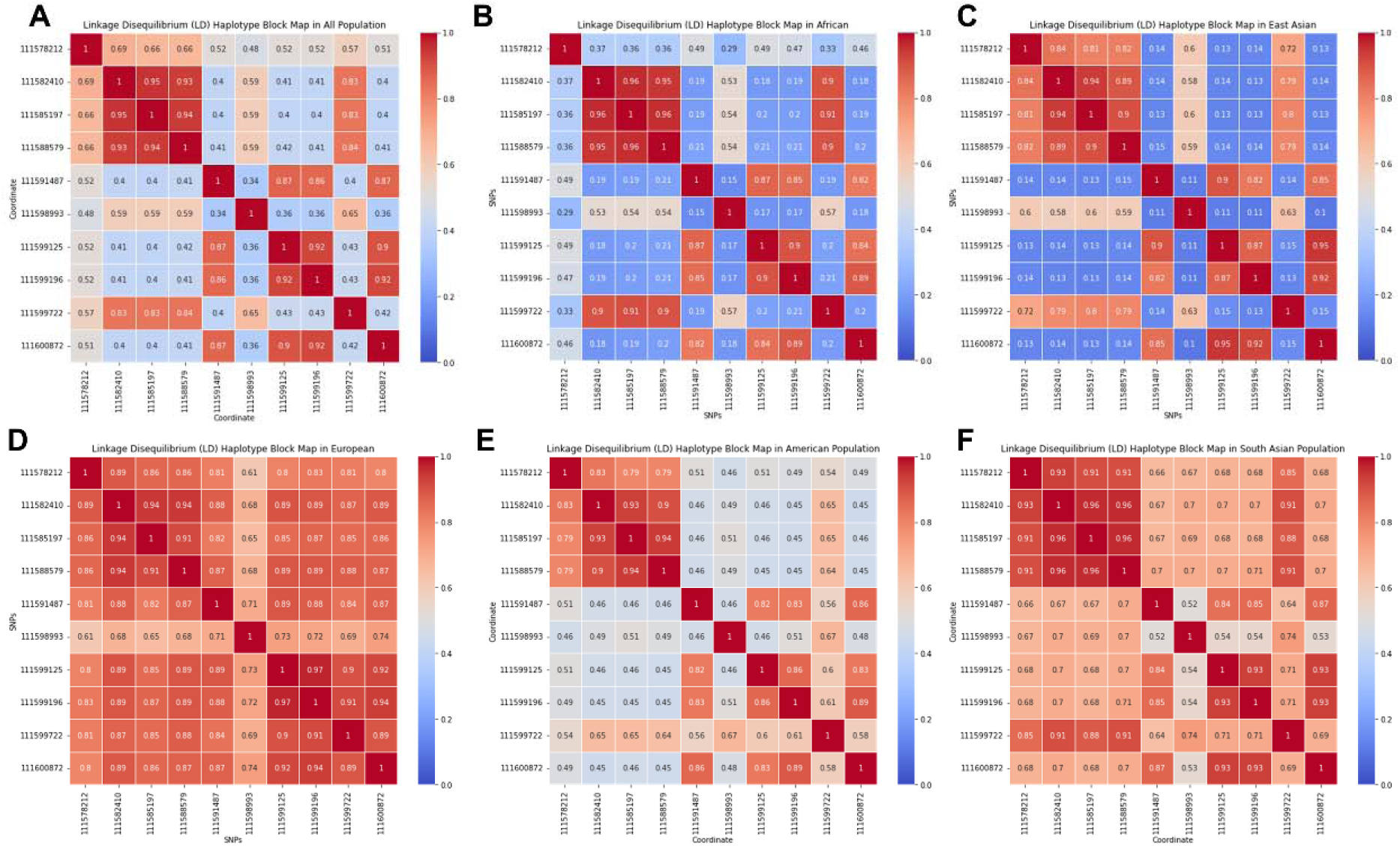
Heatmaps of linkage disequilibrium correlation coefficient of top 10 alternate alleles in different populations. Heatmaps showing LD correlation coefficient in A) All population . B) African. C) East Asian. D) European. E) American. F) South Asian

**Figure S3.**
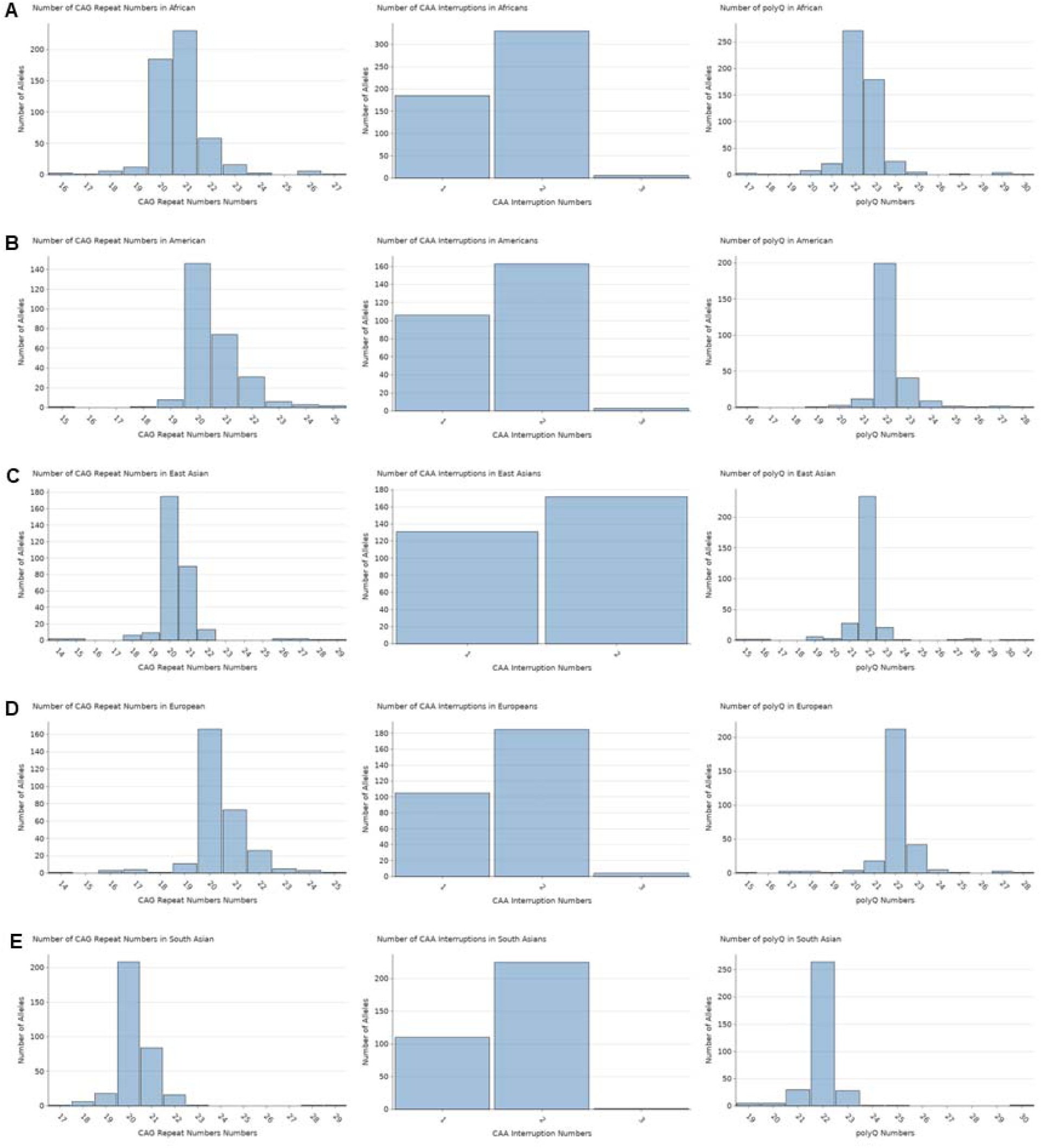
A) Histogram showing number of CAG repeats, CAA interruptions and polyQ in African population. B) Histogram showing number of CAG repeats, CAA interruptions and polyQ in American population. C) Histogram showing number of CAG repeats, CAA interruptions and polyQ in East Asian population. D) Histogram showing number of CAG repeats, CAA interruptions and polyQ in European population. E) Histogram showing number of CAG repeats, CAA interruptions and polyQ in South Asian population.

**Table S2.** Overview of haplotype frequencies. A) Haplotype frequency for haplotype at chr12: 111582410, 111585197, 111588579 and 111599722 in all population. B) Haplotype frequency for haplotype at chr12: 111582410, 111585197, 111588579 and 111599722 in population with one CAA interruption. C) Haplotype frequency for haplotype at chr12: 111582410, 111585197, 111588579 and 111599722 in population with two CAA interruptions. D) Haplotype frequency for haplotype at chr12:111591487, 111599125, 111599196 and 111600872 in all population. E) Haplotype frequency for haplotype at chr12:111591487, 111599125, 111599196 and 111600872 in population with one CAA interruption. F) Haplotype frequency for haplotype at chr12:111591487, 111599125, 111599196 and 111600872 in population with two CAA interruptions. G) Haplotype frequency at chr12:111627093, 111606529, 111599196, 111581995 and 111556082 (from Sena et. al. 2024)[25] in all long-read population dataset.

| Haplotype | All Population | African | American | East Asian | European | South Asian |
| --- | --- | --- | --- | --- | --- | --- |
| C-C-C-G | 0.700 | 0.766 | 0.651 | 0.587 | 0.712 | 0.728 |
| T-T-T-C | 0.194 | 0.165 | 0.129 | 0.284 | 0.199 | 0.209 |
| T-C-C-G | 0.047 | 0.017 | 0.151 | 0.030 | 0.027 | 0.029 |
| C-T-T-C | 0.019 | 0.019 | 0.019 | 0.040 | 0.014 | 0.0060 |
| C-C-T-G | 0.0064 | 0.0038 | 0.017 | 0.0066 | 0.010 | 0.0090 |

B
| Haplotype | All Population | African | American | East Asian | European | South Asian |
| --- | --- | --- | --- | --- | --- | --- |
| C-C-C-G | 0.313 | 0.449 | 0.226 | 0.214 | 0.298 | 0.303 |
| T-T-T-C | 0.513 | 0.459 | 0.330 | 0.618 | 0.548 | 0.624 |
| T-C-C-G | 0.088 | 0.027 | 0.349 | 0.038 | 0.048 | 0.037 |
| C-T-T-C | 0.027 | 0.011 | 0.028 | 0.069 | 0.019 | 0.0092 |
| C-C-T-G | 0.0047 | 0.0054 | 0.0094 | 0 | 0.0096 | 0.0092 |

C
| Haplotype | All Population | African | American | East Asian | European | South Asian |
| --- | --- | --- | --- | --- | --- | --- |
| C-C-C-G | 0.926 | 0.943 | 0.920 | 0.872 | 0.941 | 0.933 |
| T-T-T-C | 0.0084 | 0.0030 | 0 | 0.029 | 0.011 | 0.0089 |
| T-C-C-G | 0.020 | 0.012 | 0.025 | 0.023 | 0.016 | 0.027 |
| C-T-T-C | 0.014 | 0.024 | 0.0061 | 0.017 | 0.011 | 0.0044 |
| C-C-T-G | 0.0074 | 0.0030 | 0.0030 | 0.012 | 0.011 | 0.013 |

**Table S3.** Two-way ANOVA tests to examine the effect of interruption number and ancestry on SNVs. A) Two-way ANOVA test shows C-C-C-G haplotype depends on interruption numbers and ancestry. B) Two-way ANOVA test shows T-G-T-G haplotype depends on interruption numbers only, not based on ancestry.

D
| Haplotype | All Population | African | American | East Asian | European | South Asian |
| --- | --- | --- | --- | --- | --- | --- |
| C-C-C-A | 0.535 | 0.720 | 0.422 | 0.851 | 0.223 | 0.325 |
| T-G-T-G | 0.394 | 0.220 | 0.467 | 0.122 | 0.699 | 0.588 |
| T-G-T-A | 0.014 | 0.0077 | 0.011 | 0.0066 | 0.027 | 0.021 |
| C-C-C-G | 0.011 | 0.0095 | 0.011 | 0.0033 | 0.010 | 0.021 |
| C-G-T-G | 0.0069 | 0.011 | 0.0037 | 0 | 0.014 | 0.0030 |

E
| Haplotype | All Population | African | American | East Asian | European | South Asian |
| --- | --- | --- | --- | --- | --- | --- |
| C-C-C-A | 0.780 | 0.865 | 0.736 | 0.939 | 0.606 | 0.651 |
| T-G-T-G | 0.142 | 0.0092 | 0.160 | 0.031 | 0.260 | 0.229 |
| T-G-T-A | 0.0140 | 0.0091 | 0.018 | 0 | 0.022 | 0.022 |
| C-C-C-G | 0.0056 | 0.0091 | 0 | 0.0058 | 0.0054 | 0.0044 |
| C-G-T-G | 0.0065 | 0.0091 | 0.061 | 0 | 0.011 | 0.0044 |

F
| Haplotype | All Population | African | American | East Asian | European | South Asian |
| --- | --- | --- | --- | --- | --- | --- |
| C-C-C-A | 0.392 | 0.637 | 0.221 | 0.785 | 0.011 | 0.169 |
| T-G-T-G | 0.542 | 0.293 | 0.663 | 0.192 | 0.941 | 0.76 |
| T-G-T-A | 0.0140 | 0.0091 | 0.018 | 0 | 0.022 | 0.022 |
| C-C-C-G | 0.0056 | 0.0091 | 0 | 0.0058 | 0.0054 | 0.0044 |
| C-G-T-G | 0.0065 | 0.0091 | 0.061 | 0 | 0.011 | 0.0044 |

G
| Haplotype | All Population | African | American | East Asian | European | South Asian |
| --- | --- | --- | --- | --- | --- | --- |
| A-G-C-C-C | 0.010 | 0.0057 | 0.015 | 0.0099 | 0.0014 | 0.012 |
| G-C-G-A-T | 0.0092 | 0.0096 | 0.0074 | 0.0066 | 0.0068 | 0.015 |
| G-C-C-C-C | 0.122 | 0.358 | 0.040 | 0.013 | 0.0068 | 0.021 |
| A-C-C-C-C | 0.035 | 0.040 | 0.066 | 0.0099 | 0.027 | 0.033 |
| A-G-C-G-T | 0 | 0 | 0 | 0 | 0 | 0 |

## References

1. Ibanez K, Jadhav B, Zanovello M, Gagliardi D, Clarkson C, Facchini S, Garg P, Martin-Trujillo A, Gies SJ, Galassi Deforie V, et al: Increased frequency of repeat expansion mutations across different populations. Nat Med 2024.

2. Depienne C, Mandel JL: 30 years of repeat expansion disorders: What have we learned and what are the remaining challenges? Am J Hum Genet 2021, 108:764–785.

3. Paulson H: Repeat expansion diseases. Handb Clin Neurol 2018, 147:105–123.

4. Chen Z, Morris HR, Polke J, Wood NW, Gandhi S, Ryten M, Houlden H, Tucci A: Repeat expansion disorders. Pract Neurol 2024.

5. Logsdon GA, Vollger MR, Eichler EE: Long-read human genome sequencing and its applications. Nat Rev Genet 2020, 21:597–614.

6. Cen Z, Jiang Z, Chen Y, Zheng X, Xie F, Yang X, Lu X, Ouyang Z, Wu H, Chen S, et al: Intronic pentanucleotide TTTCA repeat insertion in the SAMD12 gene causes familial cortical myoclonic tremor with epilepsy type 1. Brain 2018, 141:2280–2288.

7. Wallenius J, Kafantari E, Jhaveri E, Gorcenco S, Ameur A, Karremo C, Dobloug S, Karrman K, de Koning T, Ilinca A, et al: Exonic trinucleotide repeat expansions in ZFHX3 cause spinocerebellar ataxia type 4: A poly-glycine disease. Am J Hum Genet 2024, 111:82–95.

8. Snyder MW, Adey A, Kitzman JO, Shendure J: Haplotype-resolved genome sequencing: experimental methods and applications. Nat Rev Genet 2015, 16:344–358.

9. Porubsky D, Garg S, Sanders AD, Korbel JO, Guryev V, Lansdorp PM, Marschall T: Dense and accurate whole-chromosome haplotyping of individual genomes. Nat Commun 2017, 8:1293.

10. Jain M, Koren S, Miga KH, Quick J, Rand AC, Sasani TA, Tyson JR, Beggs AD, Dilthey AT, Fiddes IT, et al: Nanopore sequencing and assembly of a human genome with ultra-long reads. Nat Biotechnol 2018, 36:338–345.

11. Fang L, Monteys AM, Durr A, Keiser M, Cheng C, Harapanahalli A, Gonzalez-Alegre P, Davidson BL, Wang K: Haplotyping SNPs for allele-specific gene editing of the expanded huntingtin allele using long-read sequencing. HGG Adv 2023, 4:100146.

12. Sachdev A, Gill K, Sckaff M, Birk AM, Aladesuyi Arogundade O, Brown KA, Chouhan RS, Issagholian-Lewin PO, Patel E, Watry HL, et al: Reversal of C9orf72 mutation-induced transcriptional dysregulation and pathology in cultured human neurons by allele-specific excision. Proc Natl Acad Sci U S A 2024, 121:e2307814121.

13. Monteys AM, Ebanks SA, Keiser MS, Davidson BL: CRISPR/Cas9 Editing of the Mutant Huntingtin Allele In Vitro and In Vivo. Mol Ther 2017, 25:12–23.

14. Meierhofer D, Halbach M, Sen NE, Gispert S, Auburger G: Ataxin-2 (Atxn2)- Knock-Out Mice Show Branched Chain Amino Acids and Fatty Acids Pathway Alterations. Mol Cell Proteomics 2016, 15:1728–1739.

15. Pfeffer M, Gispert S, Auburger G, Wicht H, Korf HW: Impact of Ataxin-2 knock out on circadian locomotor behavior and PER immunoreaction in the SCN of mice. Chronobiol Int 2017, 34:129–137.

16. Laffita-Mesa JM, Paucar M, Svenningsson P: Ataxin-2 gene: a powerful modulator of neurological disorders. Curr Opin Neurol 2021, 34:578–588.

17. Amado DA, Robbins AB, Whiteman KR, Smith AR, Chillon G, Chen Y, Fuller JA, Patty NA, Izda A, Cheng C, et al: AAV-based delivery of RNAi targeting ataxin-2 improves survival and pathology in TDP-43 mice. Nat Commun 2025, 16:5334.

18. Kim G, Nakayama L, Blum JA, Akiyama T, Boeynaems S, Chakraborty M, Couthouis J, Tassoni-Tsuchida E, Rodriguez CM, Bassik MC, Gitler AD: Genome-wide CRISPR screen reveals v-ATPase as a drug target to lower levels of ALS protein ataxin-2. Cell Rep 2022, 41:111508.

19. Imbert G, Saudou F, Yvert G, Devys D, Trottier Y, Garnier JM, Weber C, Mandel JL, Cancel G, Abbas N, et al: Cloning of the gene for spinocerebellar ataxia 2 reveals a locus with high sensitivity to expanded CAG/glutamine repeats. Nat Genet 1996, 14:285–291.

20. Elden AC, Kim HJ, Hart MP, Chen-Plotkin AS, Johnson BS, Fang X, Armakola M, Geser F, Greene R, Lu MM, et al: Ataxin-2 intermediate-length polyglutamine expansions are associated with increased risk for ALS. Nature 2010, 466:1069–1075.

21. Sanpei K, Takano H, Igarashi S, Sato T, Oyake M, Sasaki H, Wakisaka A, Tashiro K, Ishida Y, Ikeuchi T, et al: Identification of the spinocerebellar ataxia type 2 gene using a direct identification of repeat expansion and cloning technique, DIRECT. Nat Genet 1996, 14:277–284.

22. Findlay Black H, Wright GEB, Collins JA, Caron N, Kay C, Xia Q, Arning L, Bijlsma EK, Squitieri F, Nguyen HP, Hayden MR: Frequency of the loss of CAA interruption in the HTT CAG tract and implications for Huntington disease in the reduced penetrance range. Genet Med 2020, 22:2108–2113.

23. Sonakar AK, Shamim U, Srivastava MP, Faruq M, Srivastava AK: SCA2 in the Indian population: Unified haplotype and variable phenotypic patterns in a large case series. Parkinsonism Relat Disord 2021, 89:139–145.

24. Laffita-Mesa JM, Velazquez-Perez LC, Santos Falcon N, Cruz-Marino T, Gonzalez Zaldivar Y, Vazquez Mojena Y, Almaguer-Gotay D, Almaguer Mederos LE, Rodriguez Labrada R: Unexpanded and intermediate CAG polymorphisms at the SCA2 locus (ATXN2) in the Cuban population: evidence about the origin of expanded SCA2 alleles. Eur J Hum Genet 2012, 20:41–49.

25. Sena LS, Furtado GV, Pedroso JL, Barsottini O, Cornejo-Olivas M, Nobrega PR, Braga Neto P, Soares DMB, Vargas FR, Godeiro C, et al: Spinocerebellar ataxia type 2 has multiple ancestral origins. Parkinsonism Relat Disord 2024, 120:105985.

26. Yu Z, Zhu Y, Chen-Plotkin AS, Clay-Falcone D, McCluskey L, Elman L, Kalb RG, Trojanowski JQ, Lee VM, Van Deerlin VM, et al: PolyQ repeat expansions in ATXN2 associated with ALS are CAA interrupted repeats. PLoS One 2011, 6:e17951.

27. Toledo JB, Van Deerlin VM, Lee EB, Suh E, Baek Y, Robinson JL, Xie SX, McBride J, Wood EM, Schuck T, et al: A platform for discovery: The University of Pennsylvania Integrated Neurodegenerative Disease Biobank. Alzheimers Dement 2014, 10:477–484 e471.

28. Patterson M, Marschall T, Pisanti N, van Iersel L, Stougie L, Klau GW, Schonhuth A: WhatsHap: Weighted Haplotype Assembly for Future-Generation Sequencing Reads. J Comput Biol 2015, 22:498–509.

29. Edgar RC: MUSCLE: a multiple sequence alignment method with reduced time and space complexity. BMC Bioinformatics 2004, 5:113.

30. Dolzhenko E, Deshpande V, Schlesinger F, Krusche P, Petrovski R, Chen S, Emig-Agius D, Gross A, Narzisi G, Bowman B, et al: ExpansionHunter: a sequence-graph-based tool to analyze variation in short tandem repeat regions. Bioinformatics 2019, 35:4754–4756.

31. Cifello J, Kuksa PP, Saravanan N, Valladares O, Wang LS, Leung YY: hipFG: high-throughput harmonization and integration pipeline for functional genomics data. Bioinformatics 2023, 39.

32. Kuksa PP, Leung YY, Gangadharan P, Katanic Z, Kleidermacher L, Amlie-Wolf A, Lee CY, Qu L, Greenfest-Allen E, Valladares O, Wang LS: FILER: a framework for harmonizing and querying large-scale functional genomics knowledge. NAR Genom Bioinform 2022, 4:lqab123.

33. Consortium GT: The GTEx Consortium atlas of genetic regulatory effects across human tissues. Science 2020, 369:1318–1330.

34. Lopes KP, Snijders GJL, Humphrey J, Allan A, Sneeboer MAM, Navarro E, Schilder BM, Vialle RA, Parks M, Missall R, et al: Genetic analysis of the human microglial transcriptome across brain regions, aging and disease pathologies. Nat Genet 2022, 54:4–17.

35. Jung I, Schmitt A, Diao Y, Lee AJ, Liu T, Yang D, Tan C, Eom J, Chan M, Chee S, et al: A compendium of promoter-centered long-range chromatin interactions in the human genome. Nat Genet 2019, 51:1442–1449.

36. Teng L, He B, Wang J, Tan K: 4DGenome: a comprehensive database of chromatin interactions. Bioinformatics 2015, 31:2560–2564.

37. Nott A, Holtman IR, Coufal NG, Schlachetzki JCM, Yu M, Hu R, Han CZ, Pena M, Xiao J, Wu Y, et al: Brain cell type-specific enhancer-promoter interactome maps and disease-risk association. Science 2019, 366:1134–1139.

38. Wang Y, Song F, Zhang B, Zhang L, Xu J, Kuang D, Li D, Choudhary MNK, Li Y, Hu M, et al: The 3D Genome Browser: a web-based browser for visualizing 3D genome organization and long-range chromatin interactions. Genome Biol 2018, 19:151.

39. Gustafson JA, Gibson SB, Damaraju N, Zalusky MPG, Hoekzema K, Twesigomwe D, Yang L, Snead AA, Richmond PA, De Coster W, et al: High-coverage nanopore sequencing of samples from the 1000 Genomes Project to build a comprehensive catalog of human genetic variation. Genome Res 2024, 34:2061–2073.

40. Schloissnig S, Pani S, Rodriguez-Martin B, Ebler J, Hain C, Tsapalou V, Soylev A, Huther P, Ashraf H, Prodanov T, et al: Long-read sequencing and structural variant characterization in 1,019 samples from the 1000 Genomes Project. bioRxiv 2024.

41. Ahsan MU, Liu Q, Fang L, Wang K: NanoCaller for accurate detection of SNPs and indels in difficult-to-map regions from long-read sequencing by haplotype-aware deep neural networks. Genome Biol 2021, 22:261.

42. Pulst SM, Nechiporuk A, Nechiporuk T, Gispert S, Chen XN, Lopes-Cendes I, Pearlman S, Starkman S, Orozco-Diaz G, Lunkes A, et al: Moderate expansion of a normally biallelic trinucleotide repeat in spinocerebellar ataxia type 2. Nat Genet 1996, 14:269–276.

43. Andres AM, Lao O, Soldevila M, Calafell F, Bertranpetit J: Dynamics of CAG repeat loci revealed by the analysis of their variability. Hum Mutat 2003, 21:61–70.

44. Karczewski KJ, Francioli LC, Tiao G, Cummings BB, Alfoldi J, Wang Q, Collins RL, Laricchia KM, Ganna A, Birnbaum DP, et al: The mutational constraint spectrum quantified from variation in 141,456 humans. Nature 2020, 581:434–443.

45. Campbell MC, Tishkoff SA: African genetic diversity: implications for human demographic history, modern human origins, and complex disease mapping. Annu Rev Genomics Hum Genet 2008, 9:403–433.

46. Xu P, Pan F, Roland C, Sagui C, Weninger K: Dynamics of strand slippage in DNA hairpins formed by CAG repeats: roles of sequence parity and trinucleotide interrupts. Nucleic Acids Res 2020, 48:2232–2245.

47. Meijboom KE, Abdallah A, Fordham NP, Nagase H, Rodriguez T, Kraus C, Gendron TF, Krishnan G, Esanov R, Andrade NS, et al: CRISPR/Cas9-mediated excision of ALS/FTD-causing hexanucleotide repeat expansion in C9ORF72 rescues major disease mechanisms in vivo and in vitro. Nat Commun 2022, 13:6286.

48. Ekman FK, Ojala DS, Adil MM, Lopez PA, Schaffer DV, Gaj T: CRISPR-Cas9- Mediated Genome Editing Increases Lifespan and Improves Motor Deficits in a Huntington’s Disease Mouse Model. Mol Ther Nucleic Acids 2019, 17:829–839.

49. Hanhisuanto M, Solje E, Jokela M, Sipila JOT: Amyotrophic Lateral Sclerosis in Southwestern and Eastern Finland. Neuroepidemiology 2023, 57:238–245.

